# Persistent SARS-CoV-2 replication in severe COVID-19

**DOI:** 10.1101/2020.06.10.20127837

**Authors:** María Dolores Folgueira, Joanna Luczkowiak, Fátima Lasala, Alfredo Pérez-Rivilla, Rafael Delgado

**Author notes:** **Corresponding author:** Dr. MD Folgueira, Clinical Microbiology Department, Hospital Universitario 12 de Octubre, mail.

## Abstract

**Background:** The diagnosis of SARS-CoV-2 infection is based on viral RNA detection by real-time RT-PCR (rRT-PCR) in respiratory samples. This detection can remain positive for weeks without implying virus viability.

**Methods:** We have performed cell culture to assess viral replication in 106 respiratory samples rRT-PCR positive for SARS-CoV-2 from 105 patients with COVID-19. Fifty were samples from 50 patients with mild forms of COVID-19 who did not require hospital admission. Fifty-six samples were obtained from 55 hospitalized patients with severe pneumonia. Samples were obtained at different time points covering the time from clinical diagnosis to the follow up during hospital care.

**Results:** In 49 samples (49/106, 46.2%) a cytopathic effect (CPE) was detected in cell culture. Our study demonstrates that while in patients with mild COVID-19, viral viability is maintained in fact up to 10 days in patients with severe COVID-19 the virus can remain viable for up to 32 days after the onset of symptoms. Patients with severe COVID-19 as compared with mild cases, presented infective virus in a significantly higher proportion in samples with moderate or low viral load (Ct value > 26): 22/46 (47.8%) versus 7/38 (18.4%), (*p* <0.01), respectively.

**Conclusions:** Persistent SARS-CoV-2 replication could be demonstrated in severe COVID-19 cases for periods up to 32 days after the onset of symptoms and even at high Ct values. COVID-19 severity is a more determining factor for viral viability than the time elapsed since the onset of symptoms or the Ct value obtained in the RT-PCR assay.

## Background

COVID-19 is an acute respiratory tract infection caused by a new human coronavirus, SARS-CoV-2, that emerged in Wuhan, China, in late 2019^1,2^. On January 31st the first case of COVID-19 was detected in Spain, an imported case from Germany in Canary Islands, and thereafter on February 25th the first case was detected in Madrid ^3^. The first case of COVID-19 was confirmed at the Hospital Universitario 12 de Octubre on March 1st, a large teaching hospital with 1200 beds, covering an area over 400,000 inhabitants in southern Madrid.

The main test for the diagnosis of COVID-19 is the detection by molecular diagnostic techniques (real-time RT-PCR [rRT-PCR]) of the SARS-CoV-2 virus in respiratory samples ^4^. Detection of virus RNA is very sensitive to diagnose COVID-19 disease during the acute phase, since active replication appears to take place in the upper airway^5^. The kinetics of SARS-CoV-2 replication and detection by rRT-PCR in nasopharyngeal specimens has been well described in mild cases where viral replication is detectable up to the 8^th^ day post-symptoms ^5,6^. Detection of viral RNA by rRT-PCR is more persistent and does not necessarily indicate viable virus forms ^7^. The infection cycle of SARS-CoV-2 in severe cases of pneumonia requiring hospitalization and respiratory support is much less known. It has been suggested that the viability of the virus in respiratory samples is related to the time elapsed from the onset of symptoms and to the cycle threshold (Ct) value obtained in the amplification assay^5,8^. This value is used as a relative measure of quantification. In this study we have compared detection by rRT-PCR and the infectivity of SARS-CoV-2 in respiratory samples from mild COVID-19 cases to those from severe hospitalized cases of bilateral pneumonia.

## Methods

### Samples and patients

One hundred and six respiratory samples (102 nasopharyngeal exudates and 4 bronchial aspirates) sent to the laboratory of diagnostic virology of our institution for the detection of SARS-CoV-2 were processed both by rRT-PCR and cell culture. All samples were from adult patients. Fifty samples were obtained from 50 patients with symptoms compatible with COVID-19 who did not require hospital admission and were mostly health care workers (HCW) attending the Occupational Health and Safety Service (OHSS) for first consultation or follow up after a first positive rRT-PCR sample. Fifty-six samples were obtained from 55 hospitalized patients with severe COVID-19 pneumonia. Diagnostic of severe COVID-19 was established by respiratory, laboratory and radiographic findings. Samples were obtained at different time points covering the time from clinical diagnosis to the follow up during hospital care. Bronchial aspirates were collected during the follow up of patients admitted in the Intensive Care Unit. The study was approved by the IRB of Hospital Universitario 12 de Octubre (Reference 20-232).

### Microbiological methods

Nasopharyngeal samples were collected with flocked swabs in UTM™ viral transport medium (Copan Diagnostics, Brescia, Italy). Bronchial aspirates were diluted 1:1 in UTM™ viral transport medium upon arrival at the laboratory. Previously published rRT-PCR protocol to detect E gene^4,5^ was adapted for processing on the automated molecular diagnostic platform Panther Fusion, using its open access functionality^9^. Five hundred μl of each sample was transferred to a specimen lysis tube and loaded directly onto the Panther Fusion System. RNA extraction and amplification reagents (Open Access RNA/DNA enzyme cartridges, Extraction Reagent-S and Internal Control-S) were provided by Hologic (San Diego, CA, USA). Primers and probe for E gene were provided by TIB MolBiol (Berlin, Germany). The Ct value obtained in this assay was used as a measure of relative quantification throughout the study. For cell culture, an aliquot (250 μl) of the residual sample was decontaminated using gentamicin and amphotericin B, and inoculated in 24-well plates onto Vero E6 cells and cultured in Medium 199 supplemented with L-glutamine and 10% of fetal bovine serum. Plates were incubated in a CO_2_ 5% atmosphere for 5 days. The development of CPE was examined daily. SARS-CoV-2 CPE specificity was confirmed by immunofluorescence (shell-vial technique) with a human serum with a high titer of anti-RBD IgG as primary antibody, and a FITC-labeled anti-human IgG as secondary antibody ^10^. Additionally, upon CPE observation, culture supernatants were collected from each well, and rRT-PCR performed and confirmed to be positive at least 3 Ct lower than that of the original sample. All procedures related to cell culture were performed at a BLS3 facility.

### Data analysis

Demographic data, COVID-19 severity, symptom time to test (STT), Ct values and CPE detection were recorded and analyzed. Quantitative variables were described using median and interquartile range (IQR) and compared by Mann-Whitney U test. Categorical variables were expressed by relative frequency and compared by Fisher’s exact test. Statistics were performed on Graph Pad Prism V8 software. P-values <0.05 were considered to be statistically significant.

## Results

### Characteristics of the patients

Median age of patients with mild COVID-19 was 42 years (interquartile range (IQR): 50-32), with 78% (39/50) being women. Age and sex correspond to the profile of HCW in our setting. Hospitalized patients with severe COVID-19 had a median age of 64 years (IQR: 78-53), with 66.9 % being men (36/55).

Outpatients consulted for their symptoms at the beginning of the clinical symptoms (median: 3 days, IQR: 3-2), while severe cases attended the Emergency Department after a variable time from the onset of symptoms (median: 6 days, IQR: 10-4; *p* <.001).

Six of the patients with severe COVID-19 required admission to the Intensive Care Unit (ICU) and mechanical ventilation. A total of 5 patients died, being their median age slightly higher (70 years, *p* = 0.48)

### Characteristics of the samples

For all the patients, 65 samples were obtained at the time of clinical diagnosis and 41 during patient follow-up. The median Ct value for inpatients, severe COVID-19 cases, first samples (n= 41) was 28.8 (IQR: 32.1-26.4) and for outpatients, mild COVID-19 cases, (n= 24) 26.5 (IQR: 30.6-21.1) (*p* = 0.18). The 5 patients who died presented higher viral loads (median Ct values) than the rest of the patients with pneumonia (20.2 versus 29.7, *p* = 0.008).

In the case of the samples obtained during the follow-up (n= 41), the median time elapsed from STT was 16.5 days (IQR: 23.25-13) for outpatients and 19.6 days (IQR: 27-13) for patients with severe COVID-19 (*p* = 0.54). Regarding the Ct values obtained in the rRT-PCR, the median was 37.5 (IQR: 39.3-33-1) for outpatients (n= 26), compared to 35.4 (IQR: 38.3-27.7) for inpatients (n= 15) (*p* = 0.21).

### Cell culture

In 49 samples (49/106, 46.2%), a CPE was detected in the cell culture. CPE was detectable in most cases (43/49, 87.7%) in less than 72 hours. Eighteen (18/50, 36.0%) of the samples belonged to outpatient group, while 31 samples (31/56, 53.4 %) with viable virus came from inpatients with severe COVID-19 (*p*= 0.053). In the case of the deceased patients (n=5), all their samples were CPE positive in cell culture (*p* = 0.058)

### Correlation between virus viability and time after symptoms onset

In outpatients, CPE was detected in 70.8% (17/24) of the samples obtained in the first week after the onset of symptoms. In this group of mild COVID-19 cases the maximal STT of a CPE positive sample during follow up was 10 days. In hospitalized patients with severe COVID-19, the virus was viable in a total of 55,3% (31/56) of samples: 56% (14/25) of those obtained in the first week, in 60% (9/15) in the second week, in 60% (6/10) in the third week and in 33.3% (2/6) of samples obtained beyond the third week STT. Maximal STT of a CPE positive sample in severe COVID-19 was 32 days. The distribution of the samples analyzed according to the week of collection after symptoms onset, and the percentage of samples with CPE in cell culture in each week for both groups of patients, are shown in Figure 1.

**Figure 1.**
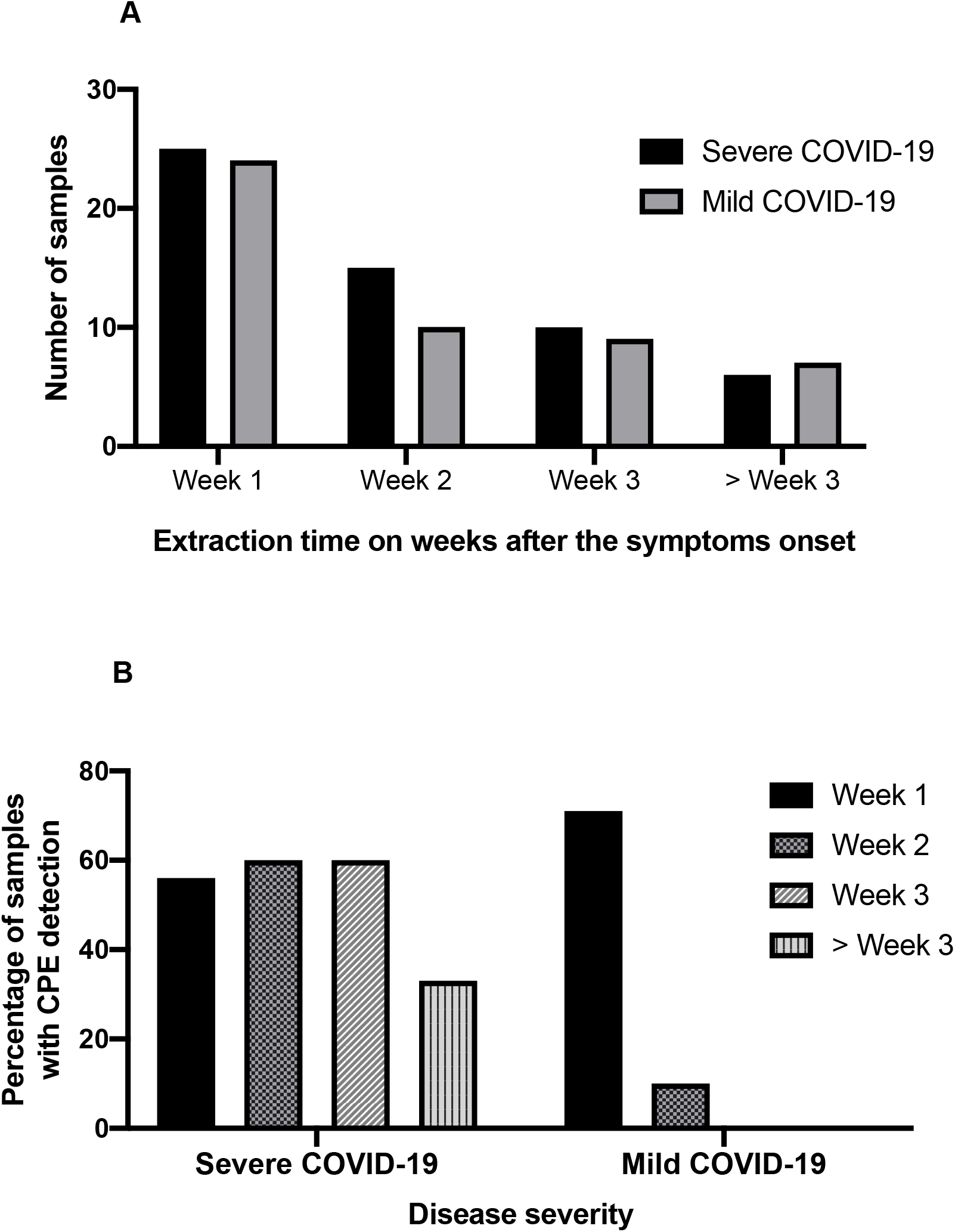
Distribution of samples according to symptom time to test (STT) on weeks (A) and percentage of samples with CPE according to STT (B) on both Severe and Mild COVID-19 groups of patients.

### Correlation between virus viability and viral load

Samples with higher viral loads (Ct value ≤ 25) in both groups of patients showed viable virus in a percentage higher than 90%. However, it is notable that even samples with low viral loads (Ct ≥ 35) could harbor viable virus, although in a much lower proportion (Figure 2). Differences in viral viability between outpatients and hospitalized patients are outstanding on samples with moderate or low viral loads (Ct ≥ 26): 22/46 (47.8%) versus 7/38 (18.4%), (*p* <0.01), respectively.

**Figure 2.**
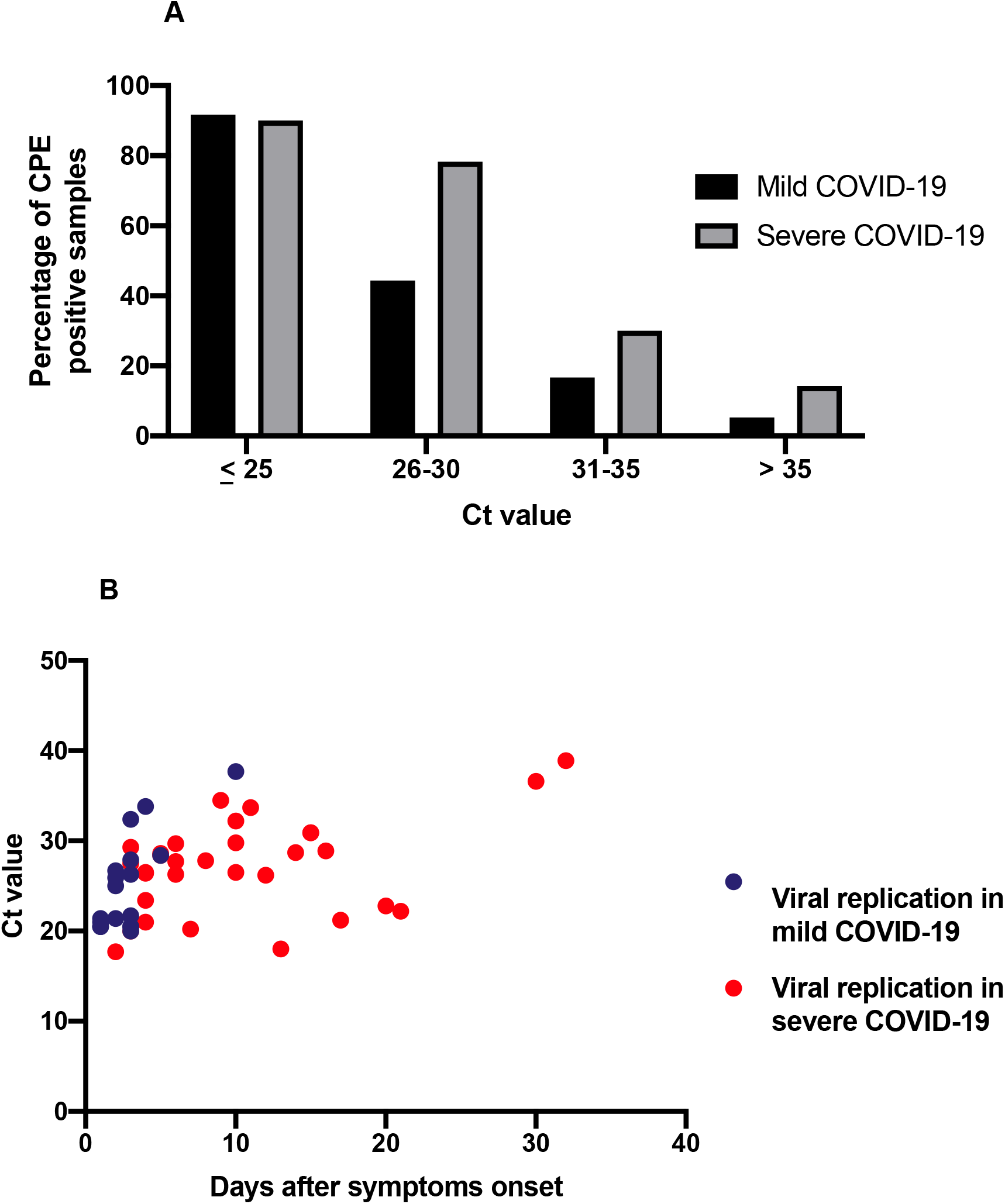
Percentage of samples with CPE according to the Ct amplification value (A) and correlation of viral replication with Ct value and symptoms onset (B) on both cohorts studied

## Discussion

The use of rRT-PCR as a diagnostic and follow-up method for SARS-CoV-2 infection has led to infer misleading information regarding the duration of infectivity of patients, the possibility of reactivation or even reinfection that has not been demonstrated so far ^7^. Although rRT-PCR is the gold standard as a diagnostic method, it is less useful as a follow-up technique, since samples from patients who have overcome both mild and severe SARS-CoV-2 infections can retain detectable viral RNA for variable periods of time^5,9,13^ Furthermore, the quantification of the virus is not well solved in respiratory samples, since their quality has a significant influence on the results. The assessment of SARS-CoV-2 viability will allow to establish criteria for the isolation of the patients, evaluation of the efficacy of the treatments used, and even prognostic value.

In severe cases of COVID-19 a hyper-inflammatory process triggered by SARS-CoV-2, typically during the second week of disease, and the subsequent expression of high levels of cytokines is invoked to explain the severe pulmonary damage leading to respiratory failure. This has led to therapeutics recommendations on using antivirals as soon as possible and manage the severe consequences of an uncontrolled immune-driven response, during the second and third week, with anti-inflammatory therapy such as steroids or IL-6 receptor antagonist monoclonal antibodies^11^. The contribution of maintained viral replication to the pathogeny of lung tissue damage and respiratory failure, along to the antiviral countermeasures, should be further explored.

Here we show a high rate (55,3%) and persistence (STT up to day 32) of positive culture in samples from severe COVI-19 cases, indicating that replication is preserved in respiratory samples in a much higher proportion than previously reported.

Our study presents the highest rate of positive culture of SARS-CoV-2 of those published to date^5,8,12^. A recent study has shown prolonged viral shedding in patients with severe COVID-19 and this fact appears to be closely related to a high viral load and to a low neutralizing antibody response^12^. Although we have observed a significant positive correlation between viral load and the presence of viable virus, the Ct value does not seem sufficient to discriminate samples harboring infective virus. We show that in patients with severe forms of COVID-19, viral replication can be detected even with moderate or low viral load during prolonged periods of time. Whether this is related to a higher cell culture positivity rate (55.3%) as compared to those reported by van Kampen JJA et al^12^ (9%) in samples from severe COVID-19 due to technical factors, such as the cell line permissiveness to SARS-CoV-2, remains to be defined.

In conclusion, we present here a large series of samples in which cell culture has been performed, including respiratory samples from patients with severe COVID-19. A high viral load indicates a greater probability of viral viability, but moderate or low viral loads cannot be used as an indicator of lack of infectivity. We have detected, as reported^5,8^, a completely different pattern of SARS-CoV-2 viability in upper respiratory tract samples from mild COVID-19 cases, where viral replication in the upper respiratory tract occurs during a short, but epidemiologically very important, period of time (maximum 10 days after symptoms onset) as compared with severe COVID-19 cases in hospitalized patients, where viable virus can be frequently demonstrated during prolonged periods up to 4 weeks both in upper and lower respiratory tract samples.

## Data Availability

All data referred to in the manuscript are available upon reasonable request

## Acknowledgements

We gratefully acknowledge M.B.A. for providing serum for culture confirmation experiments. This work was supported by grants to RD by the Instituto de Investigación Carlos III (grants FIS PI 1801007, by the European Union Commission Horizon 2020 Framework Programme: Project VIRUSCAN FETPROACT-2016: 731868. And by Fundación Caixa-Health Research (Project StopEbola) and to MDF by the Instituto de Investigación Carlos III (FIS PI1800740).

